# Racial and ethnic determinants of Covid-19 risk

**DOI:** 10.1101/2020.06.18.20134742

**Authors:** Chun-Han Lo, Long H. Nguyen, David A. Drew, Mark S. Graham, Erica T. Warner, Amit D. Joshi, Christina M. Astley, Chuan-Guo Guo, Wenjie Ma, Raaj S. Mehta, Sohee Kwon, Mingyang Song, Richard Davies, Joan Capdevila, Karla A. Lee, Mary Ni Lochlainn, Thomas Varsavsky, Carole H. Sudre, Jonathan Wolf, Yvette C. Cozier, Lynn Rosenberg, Lynne R. Wilkens, Christopher A. Haiman, Loïc Le Marchand, Julie R. Palmer, Tim D. Spector, Sebastien Ourselin, Claire J. Steves, Andrew T. Chan, on behalf of the COPE Consortium

## Abstract

**Background:** Racial and ethnic minorities have disproportionately high hospitalization rates and mortality related to the novel coronavirus disease 2019 (Covid-19). There are comparatively scant data on race and ethnicity as determinants of infection risk.

**Methods:** We used a smartphone application (beginning March 24, 2020 in the United Kingdom [U.K.] and March 29, 2020 in the United States [U.S.]) to recruit 2,414,601 participants who reported their race/ethnicity through May 25, 2020 and employed logistic regression to determine the adjusted odds ratios (aORs) and 95% confidence intervals (CIs) for a positive Covid-19 test among racial and ethnic groups.

**Results:** We documented 8,858 self-reported cases of Covid-19 among 2,259,841 non-Hispanic white; 79 among 9,615 Hispanic; 186 among 18,176 Black; 598 among 63,316 Asian; and 347 among 63,653 other racial minority participants. Compared with non-Hispanic white participants, the risk for a positive Covid-19 test was increased across racial minorities (aORs ranging from 1.24 to 3.51). After adjustment for socioeconomic indices and Covid-19 exposure risk factors, the associations (aOR [95% CI]) were attenuated but remained significant for Hispanic (1.58 [1.24-2.02]) and Black participants (2.56 [1.93-3.39]) in the U.S. and South Asian (1.52 [1.38-1.67]) and Middle Eastern participants (1.56 [1.25-1.95]) in the U.K. A higher risk of Covid-19 and seeking or receiving treatment was also observed for several racial/ethnic minority subgroups.

**Conclusions:** Our results demonstrate an increase in Covid-19 risk among racial and ethnic minorities not completely explained by other risk factors for Covid-19, comorbidities, and sociodemographic characteristics. Further research investigating these disparities are needed to inform public health measures.

## INTRODUCTION

The novel coronavirus disease 2019 (Covid-19) caused by the severe acute respiratory syndrome coronavirus 2 (SARS-CoV-2) continues to pose a tremendous threat to the global community. As of June 2020, over 6.4 million cases of Covid-19 have been documented worldwide with nearly 381,000 deaths.^1^ Prior investigations suggest that Covid-19 disproportionately impacts certain populations, including older individuals, males,^2^ and those diagnosed with obesity^3^ or other underlying health conditions.^4^ However, large-scale studies investigating potential racial or ethnic disparities in infection risk are limited.

Emerging data in the United States (U.S.) and the United Kingdom (U.K.) suggest that racial and ethnic minorities may account for an outsized proportion of Covid-19 hospitalizations and deaths.^5–8^ Although the U.S. Center for Disease Control and Prevention (CDC) recently began mandating the reporting of Covid-19 testing results according to race/ethnicity,^9^ data on the risk of testing positive for Covid-19 across a large population are lacking. Most estimates of Covid-19 risk to date are based on reports that are not uniformly collected, rely on a patchwork of information from local authorities, and do not account for other factors that could influence risk,^10^ such as comorbidities^3,11^, ability to practice social distancing,^12,13^ income disparity, poorer access to testing/care, and language and cultural barriers.^14–16^

Given the importance of understanding the determinants of health among ethnic minority groups, a comprehensive multinational investigation examining racial and ethnic disparities in risk of Covid-19 infection is urgently needed. We conducted a population-scale investigation to examine the risk of reporting a positive SARS-CoV-2 test and presenting for Covid-19 care among racial and ethnic groups in the U.S and the U.K.

## METHODS

### Real-time assessment of Covid-19 using smartphone technology

We recruited individuals from the general population in the U.S. and the U.K. using the Covid Symptom Study smartphone application (“app”) developed by Zoe Global Ltd. with scientific input from Massachusetts General Hospital and King’s College London.^17^ The app was launched in the U.K. on March 24 and in the U.S. March 29, 2020. It offers users a guided interface to report baseline demographic information and comorbidities. Users are prompted to use the application daily to allow for longitudinal, prospective collection of concomitant symptoms, health care visits, and Covid-19 test results. Study participants were recruited through general media, social media outreach, and direct invitations from the investigators of long-running prospective cohorts.^18^ At enrollment, participants provided informed consent to the use of aggregated information for research purposes and agreed to applicable privacy policies and terms of use. This research study was approved by the Partners Human Research Committee (Protocol 2020P000909) and King’s College London Ethics Committee (REMAS ID 18210, LRS-19/20-18210). This protocol is registered with ClinicalTrials.gov (NCT04331509).

### Assessment of risk factors, symptoms, testing, and/or care for Covid-19

Information collected through the application has previously been described.^17^ Briefly, at enrollment, participants were asked to provide information on demographic factors and suspected risk factors for Covid-19 (**Table 1**). On first use and daily, participants were asked if they felt physically normal, and if not, their symptoms, including the presence of fever, persistent cough, fatigue, and loss of smell/taste, among others (**Supplemental Table 2** & **Supplemental Table 3**). Participants were asked if they had been tested for Covid-19 and the results (none, negative, pending, or positive). Visits to the hospital for care were documented by participants during daily logs and were recorded if they reported being “in hospital” or “back from hospital”. Treatment was recorded if participants indicated receiving any of the following: supplemental oxygen, invasive ventilation, fluids, inhalers, or other treatment.

**Table 1a.** Baseline characteristics of study participants in the United States^a^.

|  | Race/Ethnicity<sup>b</sup> |  |  |  |  |  |
| --- | --- | --- | --- | --- | --- | --- |
|  | White, Non-Hispanic<br>(n=147325) | Hispanic/Latinx<br>(n=9251) | Black<br>(n=4977) | Asian<br>(n=6828) | More than one race/Other race<br>(n=4774) | Prefer not to say<br>(n=2044) |
| Age, year, % |  |  |  |  |  |  |
| < 25 | 7.9 | 17.1 | 9.1 | 9.0 | 21.0 | 7.7 |
| 25-34 | 8.6 | 16.0 | 7.3 | 12.3 | 12.0 | 10.0 |
| 35-44 | 12.9 | 19.3 | 10.1 | 15.8 | 15.5 | 15.3 |
| 45-54 | 14.5 | 16.5 | 18.1 | 15.7 | 14.0 | 17.0 |
| 55-64 | 20.6 | 13.6 | 24.9 | 12.0 | 14.5 | 19.8 |
| ≥ 65 | 35.6 | 17.5 | 30.4 | 35.2 | 23.0 | 30.2 |
| Male sex, % | 36.8 | 40.7 | 29.1 | 43.1 | 37.4 | 37.4 |
| BMI, kg/m<sup>2</sup>, % |  |  |  |  |  |  |
| 17-18.4 | 4.2 | 5.9 | 3.3 | 6.5 | 9.4 | 5.9 |
| 18.5-24.9 | 40.2 | 35.5 | 22.8 | 54.0 | 36.6 | 40.8 |
| 25-29.9 | 32.0 | 31.0 | 32.4 | 29.4 | 28.5 | 32.4 |
| ≥ 30 | 23.6 | 27.6 | 41.4 | 10.1 | 25.5 | 20.9 |
| Comorbidities, % |  |  |  |  |  |  |
| Diabetes | 5.7 | 6.9 | 13 | 8.8 | 6.4 | 5.0 |
| Heart disease | 6.5 | 4.4 | 5.9 | 6.5 | 5.9 | 6.2 |
| Lung disease and asthma | 12.6 | 13.3 | 14.8 | 10.1 | 16.8 | 11.5 |
| Kidney disease | 1.5 | 1.3 | 2.4 | 1.8 | 1.8 | 1.1 |
| Cancer<sup>b</sup> | 2.3 | 1.3 | 1.8 | 1.9 | 1.7 | 1.6 |
| Pregnant (% of females) | 0.5 | 0.9 | 0.2 | 0.6 | 0.5 | 0.2 |
| Medication usage, % |  |  |  |  |  |  |
| Immunosuppressants | 3.8 | 3.0 | 5.0 | 2.9 | 4.0 | 3.3 |
| Chemo/Immunotherapy | 0.5 | 0.4 | 0.6 | 0.5 | 0.5 | 0.4 |
| ACE inhibitor | 10.7 | 7.5 | 12.3 | 6.9 | 8.5 | 8.2 |
| Aspirin<sup>b</sup>/NSAIDs | 15.9 | 12.9 | 12.9 | 5.8 | 13.8 | 13.2 |
| Current smoker<sup>b</sup>, % | 5.0 | 7.4 | 8.3 | 4.0 | 7.9 | 4.5 |
| Isolation, % |  |  |  |  |  |  |
| Never left home | 6.1 | 8.9 | 9.5 | 5.6 | 6.1 | 4.7 |
| Rarely left home | 88.7 | 81.4 | 82.5 | 85.8 | 86.0 | 88.9 |
| Often left home | 5.3 | 9.8 | 8.1 | 8.6 | 7.9 | 6.5 |
| Frontline healthcare worker, % | 8.0 | 8.7 | 9.2 | 8.9 | 7.2 | 6.5 |
| Community exposure, % |  |  |  |  |  |  |
| Documented | 4.0 | 6.6 | 7.7 | 4.4 | 5.2 | 4.3 |
| Suspected | 5.3 | 6.7 | 5.0 | 4.1 | 6.8 | 5.0 |
| Population density, people per sq km<sup>c</sup> | 3823.9 [1074.9, 9670.5] | 7503.2 [2231.5, 16392.8] | 7368.3 [3078.7, 16588.6] | 7924.6 [4074.3, 17256.8] | 5416.2 [1452.5, 12761.6] | 4889.1 [1278.4, 11779.3] |
| Income and education |  |  |  |  |  |  |
| Median income, USD | 67986 [58375, 83591] | 67986 [58037, 81061] | 66793 [57200, 78777] | 82808 [67986, 89373] | 69475 [59838, 83695] | 67986 [59114, 84213] |
| % with Bachelor's degree | 35.9 [29.3, 44.8] | 33.1 [28.8, 42.2] | 34.1 [29.7, 43.2] | 34.7 [31.8, 46.3] | 34.3 [29.6, 44.8] | 35.4 [29.7, 44.8] |

Abbreviations: ACE, angiotensin converting enzyme; BMI, body mass index; km, kilometer; kg, kilogram; NSAIDs, non-steroidal anti-inflammatory drugs.
<sup>a</sup>Median [IQR] is presented for continuous variables. Frequencies and proportions are calculated based on the total number of participants with available data.
<sup>b</sup>History of cancer, aspirin use, and smoking status have been queried since launch in the U.S. and since 3/29/2020 in the U.K. Race and ethnicity questions were queried as of 4/18/2020.
<sup>c</sup>Population density was calculated from census data for each Zip Code Tabulation Area.

**Table 1b.** Baseline characteristics of study participants in the United Kingdom^a^.

|  | Race/Ethnicity<sup>b</sup> |  |  |  |  |  |  |  |  |
| --- | --- | --- | --- | --- | --- | --- | --- | --- | --- |
|  | White, non-Hispanic<br>(n=2104829) | Hispanic/Latinx<br>(n=2379) | Black<br>(n=13057) | South Asian<br>(n=46350) | Chinese<br>(n=7736) | East/Southeast Asian<br>(n=2110) | Middle Eastern<br>(n=8466) | More than one race/other race<br>(n=48908) | Prefer not to say<br>(n=8893) |
| Age, year, % |  |  |  |  |  |  |  |  |  |
| < 25 | 13.0 | 9.3 | 12.6 | 12.7 | 12.0 | 6.9 | 11.6 | 30.1 | 10.9 |
| 25-34 | 14.4 | 23.5 | 17.2 | 18.7 | 23.0 | 16.5 | 19.8 | 18.9 | 15.8 |
| 35-44 | 17.7 | 33.6 | 22.2 | 27.9 | 27.6 | 30.1 | 25.8 | 19.1 | 22.0 |
| 45-54 | 20.2 | 20.2 | 23.8 | 20.7 | 19.9 | 28.1 | 18.9 | 15.1 | 22.2 |
| 55-64 | 18.7 | 8.8 | 18.5 | 11.8 | 10.2 | 11.2 | 13.6 | 10.8 | 16.9 |
| ≥ 65 | 16.0 | 4.5 | 5.7 | 8.2 | 7.3 | 7.3 | 10.3 | 6.1 | 12.2 |
| Male sex, % | 41.3 | 36.8 | 46.1 | 46.7 | 34.8 | 25.5 | 50.8 | 41.4 | 49.5 |
| BMI, kg/m<sup>2</sup>, % |  |  |  |  |  |  |  |  |  |
| 17-18.4 | 5.6 | 5.5 | 5.9 | 7.0 | 8.3 | 6.7 | 5.1 | 12.9 | 7.5 |
| 18.5-24.9 | 41.5 | 44.2 | 29.8 | 46.3 | 64.8 | 61.0 | 41.6 | 44.0 | 40.6 |
| 25-29.9 | 31.4 | 26.8 | 32.9 | 30.7 | 20.0 | 22.5 | 32.1 | 24.7 | 30.4 |
| ≥ 30 | 21.5 | 23.4 | 31.4 | 16.1 | 7.0 | 9.8 | 21.2 | 18.4 | 21.6 |
| Comorbidities, % |  |  |  |  |  |  |  |  |  |
| Diabetes | 3.5 | 1.3 | 7.0 | 7.4 | 2.9 | 4.3 | 4.7 | 2.9 | 3.9 |
| Heart disease | 2.9 | 1.6 | 2.2 | 3.3 | 1.7 | 1.8 | 3.4 | 1.8 | 2.9 |
| Lung disease and asthma | 12.1 | 9.2 | 13.3 | 11.7 | 9.0 | 9.0 | 9.4 | 13.6 | 12.0 |
| Kidney disease | 0.7 | 0.8 | 1.1 | 0.9 | 0.7 | 0.5 | 1.1 | 0.7 | 0.7 |
| Cancer<sup>b</sup> | 1.4 | 1.0 | 1.4 | 0.8 | 0.9 | 1.4 | 1.1 | 0.8 | 1.4 |
| Pregnant (% of females) | 1.0 | 1.2 | 0.8 | 1.5 | 1.3 | 1.3 | 1.5 | 1.1 | 1.0 |
| Medication usage, % |  |  |  |  |  |  |  |  |  |
| Immunosuppressants | 3.4 | 3.0 | 4.0 | 3.6 | 2.2 | 2.6 | 2.9 | 3.3 | 3.7 |
| Chemo/Immunotherapy | 0.3 | 0.3 | 0.2 | 0.2 | 0.3 | 0.3 | 0.3 | 0.2 | 0.3 |
| ACE inhibitor | 6.0 | 2.0 | 5.9 | 4.8 | 2.6 | 3.4 | 4.3 | 3.2 | 4.8 |
| Aspirin<sup>b</sup>/NSAIDs | 5.9 | 6.9 | 7.0 | 4.0 | 2.5 | 3.6 | 5.6 | 5.2 | 5.8 |
| Current smoker<sup>b</sup>, % | 8.3 | 8.5 | 11.2 | 7.9 | 5.3 | 7.7 | 14.9 | 10.2 | 10.8 |
| Isolation, % |  |  |  |  |  |  |  |  |  |
| Never left home | 5.7 | 4.8 | 9.8 | 6.2 | 3.4 | 7.5 | 5.8 | 4.8 | 4.0 |
| Rarely left home | 88.0 | 86.1 | 80.2 | 81.6 | 87.4 | 83.5 | 83.2 | 86.2 | 89.1 |
| Often left home | 6.3 | 9.0 | 10.0 | 12.2 | 9.2 | 9.0 | 10.9 | 8.9 | 6.9 |
| Frontline healthcare worker, % | 4.7 | 3.9 | 10.9 | 8.5 | 5.9 | 11 | 6.1 | 4.5 | 4.5 |
| Community exposure, % |  |  |  |  |  |  |  |  |  |
| Documented | 3.1 | 4.2 | 7.3 | 6.1 | 4.9 | 9.3 | 5.1 | 3.6 | 3.7 |
| Suspected | 9.1 | 14.8 | 10.4 | 9.5 | 8.3 | 11.5 | 10.4 | 12.4 | 8.3 |
| Population density, people per sq km<sup>c</sup> | 4512.0 [805.8, 10223.8] | 15448.7 [6785.6, 28932.4] | 12702.3 [6892.3, 22592.3] | 10558.1 [5097.9, 17620.7] | 11442.4 [4906.9, 22290.2] | 13270.3 [5754.3, 24413.8] | 11939.2 [5613.4, 22247.3] | 9243.5 [3186.9, 18059.9] | 7730.5 [1930.8, 15917.3] |
| Income and education |  |  |  |  |  |  |  |  |  |
| Income deprivation | 7 [5, 9] | 6 [4, 8] | 4 [3, 7] | 6 [4, 8] | 7 [4, 9] | 6 [4, 8] | 6 [4, 8.25] | 6 [4, 9] | 6 [4, 9] |
| Education deprivation | 7 [5, 9] | 8 [5, 9] | 6 [4, 8] | 7 [5, 9] | 8 [6, 9] | 7 [5, 9] | 8 [5, 9] | 7 [5, 9] | 7 [5, 9] |

Abbreviations: ACE, angiotensin converting enzyme; BMI, body mass index; km, kilometer; kg, kilogram; NSAIDs, non-steroidal anti-inflammatory drugs.
<sup>a</sup>Median [IQR] is presented for continuous variables. Frequencies and proportions are calculated based on the total number of participants with available data.
<sup>b</sup>History of cancer, aspirin use, and smoking status have been queried since 3/29/2020 in the U.K. Race and ethnicity questions were queried as of 4/18/2020.
<sup>c</sup>Population density was calculated from census data for each Lower Layer Super Output Area.

### Assessment of race and ethnicity

Individuals were asked to report with which race and/or ethnicity they self-identified. Questions were based on standard categories from the U.S. National Institutes of Health (NIH)^19^ and the U.K. Office for National Statistics.^20^ (**Supplemental Table 1**). Individuals who identified their race or ethnicity as “Other” were provided an option to enter a free-text description. Additional categories created for the U.K. include “East/Southeast Asian” and “Hispanic/Latinx”. Individuals who identified as “Mixed Race” or selected more than one race were described as “More than one race” and grouped with “Other”. We excluded participants who did not provide information on racial or ethnic identity or selected “prefer not to say”. In pooled analyses, categories in the U.S. and the U.K. were harmonized according to NIH categories.^19^ “White” and “Hispanic/Latinx” were categorized separately and those who were “Non-Hispanic white” were the referent group for all analyses. “Native Hawaiian and Pacific Islanders’’, “South Asian”, “Chinese”, and “East/Southeast Asian” were grouped as “Asian”, while “American Indian or Alaskan Natives” and “Middle Eastern” was categorized as “Other” (**Supplemental Methods)**.

### Socioeconomic and population density factors

Population density was calculated from census data for each Zip Code Tabulation Area (ZCTA) in the U.S. and Lower Layer Super Output Area (LSOA) in the U.K. In the U.S., we obtained indicators of income and education according to Zip Code from the U.S. Department of Agriculture.^21^ Comparable socioeconomic indices were quantified using the indices of multiple deprivation (IMD) at the LSOA level in the U.K.,^22^ for which we combined scores from each of the U.K.’s 4 constituent countries into a unified scale by assuming the same distribution of deprivation scores in each country.

### Statistical analysis

We employed logistic regression models to examine the odds ratios (ORs) and 95% confidence intervals (CIs) of a positive Covid-19 test and of seeking and receiving treatment. Multivariable models were conditioned upon age, date, and country (if pooled). Additional covariates were selected *a priori* based on putative risk factors, including sex, body mass index, history of diabetes, heart, lung, or kidney disease, current smoking status, isolation, community interaction with individuals with Covid-19, frontline healthcare worker status, population density, income, and education. Missing categorical data were included as a missing indicator. To account for likelihood to receive testing, we performed separate inverse probability weighting (IPW) in the U.S. and the U.K. as a function of race/ethnicity and other factors, such as age, symptom burden, COVID-19 exposure risk factors, and socioeconomic status, followed by inverse probability weighted logistic regression (**Supplemental Methods**).

In addition to a positive Covid-19 test, we utilized a previously developed symptom-based classifier predictive of positive Covid-19 testing.^23^ Briefly, using logistic regression and symptoms preceding testing, we found that a weighted score including loss of smell/taste, fatigue, persistent cough, and loss of appetite predicts Covid-19 positivity with high specificity (**Supplemental Methods**). To further examine the risk of more severe Covid-19, we evaluated the likelihood of Covid-19 and hospital visit or treatment. We conducted analyses in the U.S. and the U.K. separately but also leveraged harmonized data for pooled results. Two-sided p-values <0.05 were considered statistically significant. All analyses were performed using R 3.6.1 (Vienna, Austria).

## RESULTS

### Study population

Between March 24 and May 25, 2020, 2,414,601 participants (U.S. *n* = 179,873; U.K. *n* = 2,234,728) had registered and responded to questions within the app, including 2,259,841 non-Hispanic white, 9,615 Hispanic, 18,176 Black, 63,316 Asian participants, and 63,653 participants of more than one race/other race. Median age was 47 years (interquartile range [IQR] 33-60). Black and Hispanic participants were more likely to be overweight or obese and have diabetes and kidney disease (**Table 1**). All racial and ethnic minorities reported a lower level of social isolation, were comparatively overrepresented among frontline healthcare workers, and reported a higher likelihood of exposure to individuals with suspected or documented Covid-19. Non-Hispanic white participants more commonly reported living in less densely populated regions, and Black individuals tended to live in locales characterized by lower income and educational attainment. The prevalence of symptoms appeared largely consistent across groups (**Supplemental Table 2** & **Supplemental Table 3**). The most common symptoms reported were headache, fatigue, and sore throat.

### Risk of a positive Covid-19 test according to race and ethnicity

Among 2,414,601 adults, we documented 10,051 reports of positive Covid-19 testing. In the U.S., all racial minorities had an increased risk of reporting a positive Covid-19 test (age-adjusted ORs ranging from 1.52 to 3.69), with the highest risk among Black participants (**Table 2a**). In the U.K., increased risk was observed among Black, South Asian, East/Southeast Asian, Middle Eastern individuals, and those reporting more than one or other race (age-adjusted ORs ranging from 1.23 to 2.85) (**Table 2b**). Results were essentially unchanged with additional adjustment for comorbidities and lifestyle factors. We next performed IPW to account for the likelihood of receiving a test and found modest attenuation of the estimates, with most racial and ethnic minorities remaining significantly associated with a positive test for Covid-19.

**Table 2.** Risk of a positive Covid-19 test according to race and ethnicity in the United States and the United Kingdom adjusted for comorbidities.

| a. United States | Race/Ethnicity |  |  |  |  |  |  |  |
| --- | --- | --- | --- | --- | --- | --- | --- | --- |
|  | White, non-Hispanic | Hispanic/Latinx | Black | Asian | More than one/other race |  |  |  |
| Individuals testing positive / n | 498/147325 | 89/9251 | 65/4977 | 41/6828 | 23/4774 |  |  |  |
| Age-adjusted OR (95% CI)<sup>a</sup> | 1 (reference) | 2.69 (2.14-3.39) | 3.69 (2.83-4.81) | 1.87 (1.36-2.58) | 1.52 (1.00-2.31) |  |  |  |
| Multivariable-adjusted OR (95% CI)<sup>b</sup> | 1 (reference) | 2.68 (2.13-3.38) | 3.51 (2.68-4.60) | 1.97 (1.43-2.73) | 1.51 (0.99-2.30) |  |  |  |
| Multivariable-adjusted OR (95% CI) weighted by IPW<sup>b</sup> | 1 (reference) | 1.66 (1.18-2.34) | 2.49 (1.68-3.69) | 1.42 (0.86-2.35) | 1.32 (0.67-2.61) |  |  |  |
| b. United Kingdom | Race/Ethnicity |  |  |  |  |  |  |  |
|  | White, non-Hispanic | Hispanic/Latinx | Black | South Asian | Chinese | East/Southeast Asian | Middle Eastern | More than one/other race |
| Individuals testing positive / n | 8335/2104829 | 15/2379 | 121/13057 | 485/46350 | 44/7736 | 27/2110 | 82/8466 | 226/48908 |
| Age-adjusted OR (95% CI)<sup>a</sup> | 1 (reference) | 1.42 (0.86-2.36) | 2.17 (1.81-2.60) | 2.44 (2.23-2.68) | 1.30 (0.97-1.75) | 2.85 (1.95-4.16) | 2.28 (1.83-2.83) | 1.23 (1.08-1.40) |
| Multivariable-adjusted OR (95% CI)<sup>b</sup> | 1 (reference) | 1.41 (0.85-2.34) | 2.10 (1.75-2.51) | 2.50 (2.28-2.74) | 1.39 (1.03-1.87) | 2.93 (2.01-4.28) | 2.38 (1.91-2.96) | 1.24 (1.09-1.41) |
| Multivariable-adjusted OR (95% CI) weighted by IPW<sup>b</sup> | 1 (reference) | 1.71 (0.89-3.27) | 1.97 (1.47-2.64) | 1.68 (1.43-1.97) | 1.79 (1.08-2.96) | 1.02 (0.55-1.87) | 2.10 (1.52-1.87) | 2.10 (1.52-2.91) |

Abbreviations: CI, confidence interval; OR, odds ratio.
<sup>a</sup>Stratified by age and date of entry into the study.
<sup>b</sup>Adjusted for sex, history of diabetes, heart disease, lung disease, kidney disease, and current smoker status (each yes/no), and body mass index (17-18.4, 18.5-24.9, 25-29.9, and $\geq 30$ kg/m<sup>2</sup>).

Next, we considered the possibility that other social determinants mediated the association of race and ethnicity with risk of Covid-19 by adjusting for income and education, population density, level of isolation, frontline healthcare worker status, and community exposure to a Covid-19-positive individual (**Figure 1**). Overall, the associations (aOR [95% CI]) were moderately attenuated but remained significant for Hispanic (1.58 [1.24-2.02]) and Black users (2.56 [1.93-3.39]) in the U.S. and South Asian (1.52 [1.38-1.67]) and Middle Eastern populations (1.56 [1.25-1.95]) in the U.K.

**Figure 1.**
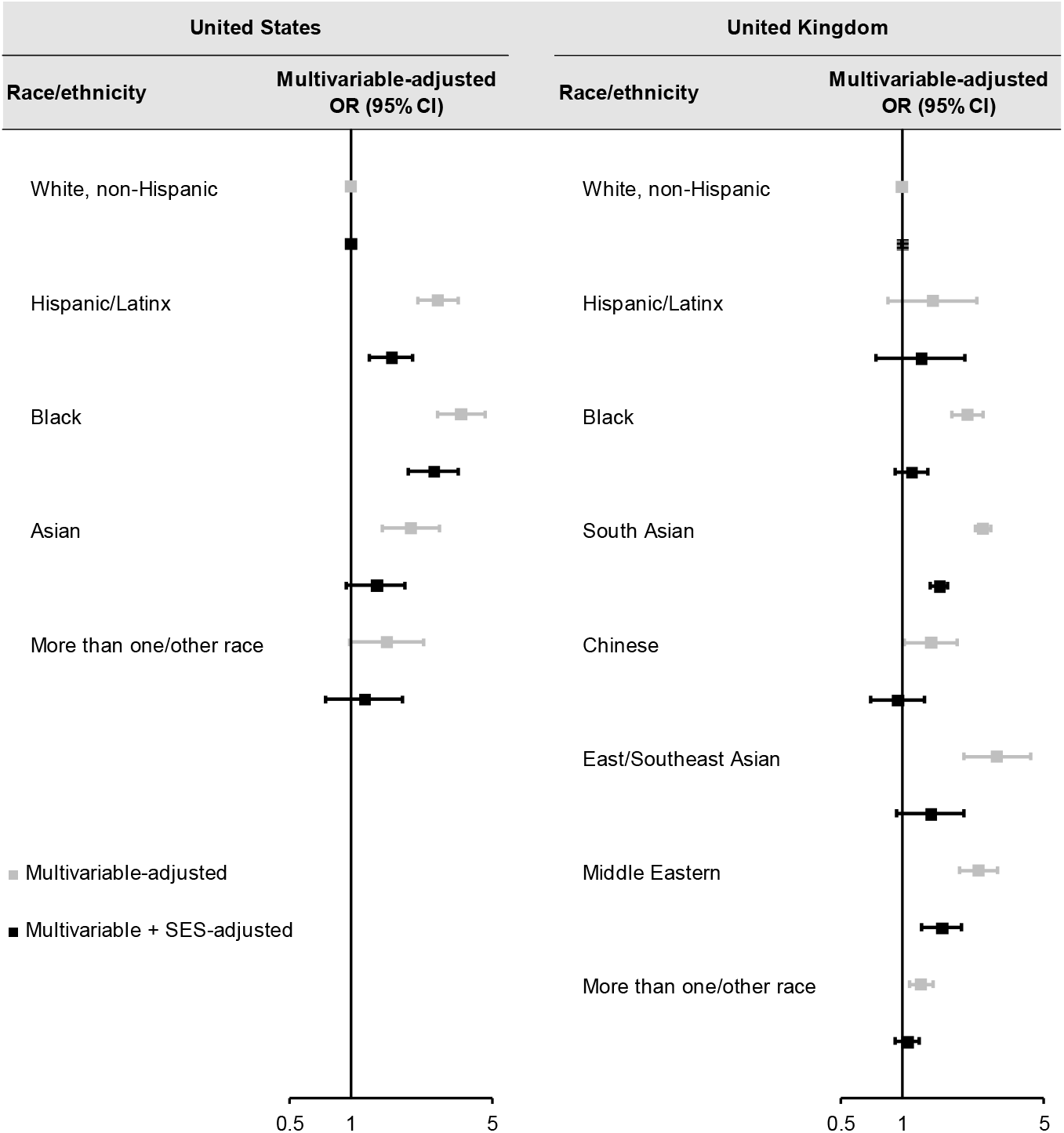
Risk of a positive Covid-19 test according to race and ethnicity in the United States and the United Kingdom with additional adjustment for socioeconomic indices. The multivariable association adjusted for comorbidities, as in Table 2, of race and ethnicity with risk of testing Covid-19 positive in each country is presented (gray). Additional adjustment for isolation (never left home, rarely left home, often left home), frontline healthcare worker (yes/no), community exposure (no, documented, suspected), population density, income, and education in each country, as described in Table 1, demonstrates attenuation of most associations.

### Risk of predicted Covid-19 using a symptom-based classifier

To further address the possibility of disparities in access to testing, we used a validated symptom-based model associated with predicted Covid-19 infection in a pooled analysis. Compared to non-Hispanic white participants, we found a significant increase in risk for predicted Covid-19 in Black (aOR 1.17 [95% CI 1.10-1.25]), Hispanic (1.11 [1.00-1.23]), Asian (1.06 [1.03-1.10]), and more than one/other racial minorities (1.21 [1.17-1.25]) (**Table 3**). Within each country, we observed similar estimates for combined racial and ethnic minority groups compared to non-Hispanic whites in the U.S. (1.20 [1.11-1.30]) and the U.K. (1.22 [1.19-1.25]) (data not shown).

**Table 3.**
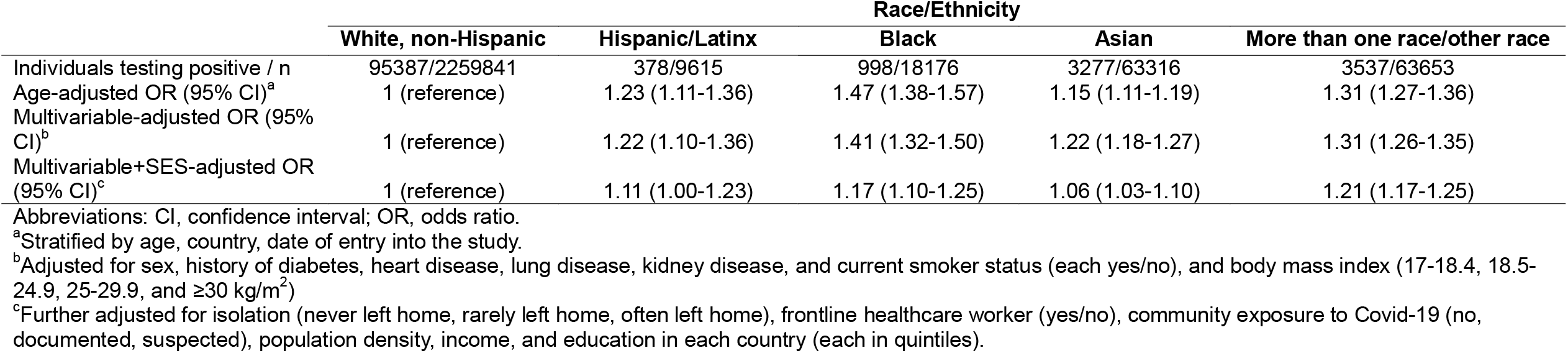
Risk of predicted Covid-19 based on a symptom score according to race and ethnicity in the United States and United Kingdom (pooled).

|  | Race/Ethnicity |  |  |  |  |
| --- | --- | --- | --- | --- | --- |
|  | White, non-Hispanic | Hispanic/Latinx | Black | Asian | More than one race/other race |
| Individuals testing positive / n | 95387/2259841 | 378/9615 | 998/18176 | 3277/63316 | 3537/63653 |
| Age-adjusted OR (95% CI)<sup>a</sup> | 1 (reference) | 1.23 (1.11-1.36) | 1.47 (1.38-1.57) | 1.15 (1.11-1.19) | 1.31 (1.27-1.36) |
| Multivariable-adjusted OR (95% CI)<sup>b</sup> | 1 (reference) | 1.22 (1.10-1.36) | 1.41 (1.32-1.50) | 1.22 (1.18-1.27) | 1.31 (1.26-1.35) |
| Multivariable+SES-adjusted OR (95% CI)<sup>c</sup> | 1 (reference) | 1.11 (1.00-1.23) | 1.17 (1.10-1.25) | 1.06 (1.03-1.10) | 1.21 (1.17-1.25) |

Abbreviations: CI, confidence interval; OR, odds ratio.
<sup>a</sup>Stratified by age, country, date of entry into the study.
<sup>b</sup>Adjusted for sex, history of diabetes, heart disease, lung disease, kidney disease, and current smoker status (each yes/no), and body mass index (17-18.4, 18.5-24.9, 25-29.9, and $\geq 30$ kg/m<sup>2</sup>)
<sup>c</sup>Further adjusted for isolation (never left home, rarely left home, often left home), frontline healthcare worker (yes/no), community exposure to Covid-19 (no, documented, suspected), population density, income, and education in each country (each in quintiles).

### Risk of Covid-19, seeking, or receiving treatment

In both the U.S. and the U.K, compared to non-Hispanic white participants, the aORs (95% CI) for Covid-19 requiring a hospital visit were elevated across racial and ethnic minorities, ranging from 1.32 (1.03-1.68) for more than one/other race to 1.64 (1.20-2.23) for Black race (**Table 4**). The corresponding aORs (95% CI) for receiving treatment were also increased for Black (1.83 [1.17-2.85]) and Asian participants (1.60 [1.19-2.14]). Country-specific estimates for combined minority groups compared to non-Hispanic whites were similar to the pooled analysis (**Supplemental Table 4** & **Supplemental Table 5**).

**Table 4.**
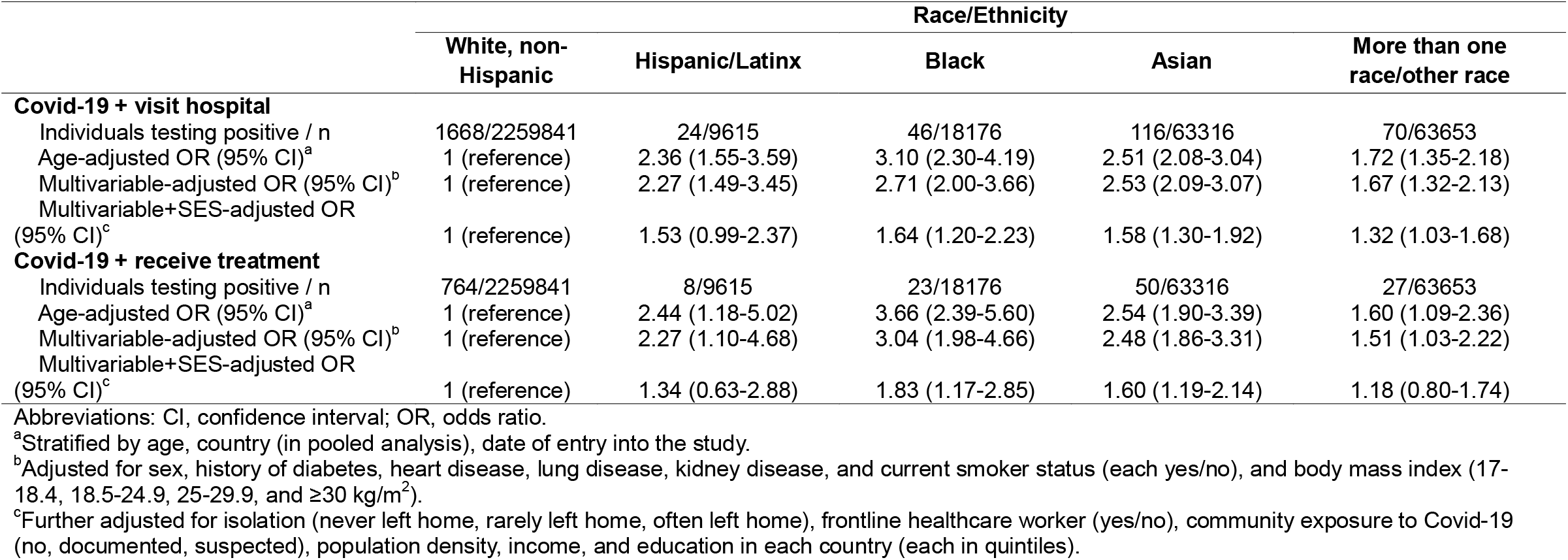
Risk of a positive COVID-19 test and seeking or receiving treatment according to race and ethnicity in the United States and the United Kingdom.

|  | Race/Ethnicity |  |  |  |  |
| --- | --- | --- | --- | --- | --- |
|  | White, non-Hispanic | Hispanic/Latinx | Black | Asian | More than one race/other race |
| **Covid-19 + visit hospital** |  |  |  |  |  |
| Individuals testing positive / n | 1668/2259841 | 24/9615 | 46/18176 | 116/63316 | 70/63653 |
| Age-adjusted OR (95% CI)<sup>a</sup> | 1 (reference) | 2.36 (1.55-3.59) | 3.10 (2.30-4.19) | 2.51 (2.08-3.04) | 1.72 (1.35-2.18) |
| Multivariable-adjusted OR (95% CI)<sup>b</sup> | 1 (reference) | 2.27 (1.49-3.45) | 2.71 (2.00-3.66) | 2.53 (2.09-3.07) | 1.67 (1.32-2.13) |
| Multivariable+SES-adjusted OR (95% CI)<sup>c</sup> | 1 (reference) | 1.53 (0.99-2.37) | 1.64 (1.20-2.23) | 1.58 (1.30-1.92) | 1.32 (1.03-1.68) |
| **Covid-19 + receive treatment** |  |  |  |  |  |
| Individuals testing positive / n | 764/2259841 | 8/9615 | 23/18176 | 50/63316 | 27/63653 |
| Age-adjusted OR (95% CI)<sup>a</sup> | 1 (reference) | 2.44 (1.18-5.02) | 3.66 (2.39-5.60) | 2.54 (1.90-3.39) | 1.60 (1.09-2.36) |
| Multivariable-adjusted OR (95% CI)<sup>b</sup> | 1 (reference) | 2.27 (1.10-4.68) | 3.04 (1.98-4.66) | 2.48 (1.86-3.31) | 1.51 (1.03-2.22) |
| Multivariable+SES-adjusted OR (95% CI)<sup>c</sup> | 1 (reference) | 1.34 (0.63-2.88) | 1.83 (1.17-2.85) | 1.60 (1.19-2.14) | 1.18 (0.80-1.74) |

Abbreviations: CI, confidence interval; OR, odds ratio.
<sup>a</sup>Stratified by age, country (in pooled analysis), date of entry into the study.
<sup>b</sup>Adjusted for sex, history of diabetes, heart disease, lung disease, kidney disease, and current smoker status (each yes/no), and body mass index (17-18.4, 18.5-24.9, 25-29.9, and $\geq 30$ kg/m<sup>2</sup>).
<sup>c</sup>Further adjusted for isolation (never left home, rarely left home, often left home), frontline healthcare worker (yes/no), community exposure to Covid-19 (no, documented, suspected), population density, income, and education in each country (each in quintiles).

## DISCUSSION

Among 2,414,601 participants, we observed that ethnic and racial minorities, particularly Black and Hispanic individuals in the U.S. and Asian and Middle Eastern individuals in the U.K., had a greater risk of testing positive for Covid-19 compared to non-Hispanic white participants. Results were essentially unchanged after adjusting for comorbid conditions. While further adjustment for socioeconomic indices and Covid-19 risk factors attenuated the associations, the risks remained significantly elevated for these minorities. These results were consistent after adjusting for the likelihood of being tested or when examining predicted Covid-19 infection based on prevalent symptoms. Racial minorities also had a modestly increased risk of Covid-19 and seeking hospital-based evaluation/treatment.

The racial and ethnic disparities reported here are consistent with studies documenting racial and ethnic differences in Covid-19 outcomes.^5,7,24^ Our data provide further support of research from the U.S. state of Louisiana showing that Black race was associated with a nearly 2-fold increased odds of hospital admission among Covid-19 patients after adjusting for comorbidities and sociodemographic factors.^24^ Similarly, a study from Public Health England demonstrated that the highest age-standardized death rates in confirmed Covid-19 cases were among people of Black ethnic groups, followed by Asian and mixed ethnic groups in the U.K., even after accounting for demographics, social deprivation, and region^7^. Although these data support a consistently higher risk of worsened Covid-19 outcomes among racial and ethnic minorities, whether the risk of infection also differs by race and ethnicity among the general community warrants further research. Increasing reports from the U.S. CDC, local state health departments,^25,26^ and numerous governmental agencies in Europe suggest a greater risk of Covid-19 among communities of color,^7^ but do not generally account for other factors that could influence the risk of infection.^10^ Our study complements these early reports, and by accounting for Covid-19 exposure risk factors, sociodemographic information, and comorbidities, our investigation better defines the magnitude of increased risk among ethnic and racial minorities compared to non-Hispanic white participants.

Our results demonstrate that comorbid conditions do not explain the increased likelihood to test positive for Covid-19 among minority populations, especially in the U.S., highlighting the considerable role of structural inequalities in elevating risk. Communities of color may be less able to effectively practice social distancing,^27^ given prior literature suggesting they are highly represented among the essential workforce^28^ and live in neighborhoods with the higher SARS-Cov-2 infection rates.^29^ We were able to show some attenuation of Covid-19 risk when accounting for income, education, population density, measures of social isolation, occupation as a healthcare worker, and exposure to a community member with Covid-19. However, after adjustment, the risk of a positive Covid-19 test remained significant for several racial and ethnic minorities, which is likely due to additional contributing factors for which we were unable to account, including insurance coverage, access to healthcare, use of public transit, and other essential occupations not specifically queried. Asian and Hispanic populations are also more likely than non-Hispanic whites to live in multigenerational households,^30^ and, like Black populations, are more likely to live in densely populated urban areas.^31^ Moreover, in the U.S. due to residential segregation, racial and ethnic minorities may live in predominantly minority neighborhoods with higher prevalence of infection, thereby increasing their risk of coming into contact with infected members of the community.^29^ Furthermore, Asian and Hispanic populations represent a higher proportion of foreign-born individuals,^30^ which poses additional challenges associated with cultural and language barriers, misinformation, immigration-related fear, and anxiety related to accessing care.^32,33^ Finally, “weathering” or chronic stress related to structural racism contributes to accelerated aging and chronic diseases that may contribute to COVID-19 risk.^34^

The strengths of this study include the use of a smartphone application to rapidly collect prospective data from a large multinational and multiethnic cohort, which offers real-time, actionable Covid-19 risk estimates to inform the public health response to an ongoing pandemic. Second, our study design examined documented, self-reported Covid-19 cases in the general smartphone user population, overcoming limitations related to capturing only more severe cases through administrative hospitalization records or death reports. Third, we examined the risk of predicted Covid-19 according to racial and ethnic groups and found results largely consistent with those of self-reported Covid-19.

This approach was not dependent on differences in testing availability which might vary across racial and ethnic groups. Finally, we collected information on and adjusted for a wide range of known or suspected risk factors for Covid-19, which are generally not available in existing registries or population-scale surveillance efforts.

Our study has several limitations. While the use of syndromic surveillance to better understand Covid-19 disparities has great strengths in flexibility, speed and sample size, this methodology is largely dependent upon self-reported data, and therefore susceptible to measurement bias, residual confounding bias, and selection (collider) bias. The probability of app participation, reporting, or access may be differential according to Covid-19 outcomes, minority status and/or covariates.^35,36^ Smartphone-based tools may preclude participation of certain populations such as elderly, low-income, or non-English speakers. Although our study had a relatively small proportion of racial-ethnic minorities compared to census estimates, we enrolled a high absolute number of individuals for most racial ethnic groups, with the notable exception of Native Americans. We attempted to mitigate selection bias through IPW and confounding through multivariable adjustment. Measurement errors in self-reported participant characteristics are possible. Future research may validate some risk factors in longitudinal studies linked to the Covid Symptom Study (e.g. Nurses’ Health Study). Finally, variables in the app were limited in scope to optimize participation. We acknowledge that the assessed racial and ethnic groups may be oversimplifications, and do not completely characterize the true heterogeneity in how participants experience race and ethnicity.

In conclusion, within a large population-based sample of individuals in the U.S. and the U.K., we demonstrate a significantly increased risk of Covid-19 and hospital-based evaluation/treatment for Covid-19 among several racial and ethnic minorities compared to non-Hispanic white individuals, which was partially explained by risk factors for exposure and traditional sociodemographic factors. Our results confirm the comparatively outsized burden of Covid-19 on ethnic and racial minorities and the need for further research to understand the basis for these health inequalities.

## Data Availability

Data collected in the app are being shared with other health researchers through the NHS-funded Health Data Research UK (HDRUK)/SAIL consortium, housed in the UK Secure e-Research Platform (UKSeRP) in Swansea. Anonymized data collected by the Covid Symptom Study app can be shared with bonafide researchers via HDRUK, provided the request is made according to their protocols and is in the public interest (see https://healthdatagateway.org/detail/9b604483-9cdc-41b2-b82c-14ee3dd705f6). US investigators are encouraged to coordinate data requests through the COPE Consortium (www.monganinstitute.org/cope-consortium). Data updates can be found at https://covid.joinzoe.com.

https://www.monganinstitute.org/cope-consortium

https://covid.joinzoe.com

## ACKNOWLEDGEMENTS

We express our sincere thanks to all of the participants who entered data into the app, including study volunteers enrolled in cohorts within the Coronavirus Pandemic Epidemiology (COPE) consortium. We thank the staff of Zoe Global Ltd., the Department of Twin Research at King’s College London, and the Clinical and Translational Epidemiology Unit at Massachusetts General Hospital for their tireless work.

